# Knowledge, Attitudes, and Practices for Tuberculosis in Ngäbe-Buglé Populations in Rural Panama

**DOI:** 10.1101/2020.09.21.20199232

**Authors:** Laura R Cuevas, Kaosoluchi Enendu, Sophia Raefsky, Param Bhatter, Emily Frisch, Louie Cao, Austin Crochetiere, Emilie Chow

## Abstract

**Background:** Panama has a high incidence of tuberculosis (TB), especially in rural communities. A patient survey was administered to assess practices, knowledge and attitudes towards TB within the Ngäbe-Buglé population.

**Methods:** A cross-sectional survey was distributed at Floating Doctors clinics. Subjects with familiarity of TB were assessed through questions about transmission, cause, symptoms, and treatment.

**Results:** Of the 106 patients who completed the TB practices survey, 68 patients (64%) knew of TB and completed the entire survey. Of the 64% who knew of TB, 61% knew medicine treated TB and over 80% could identify symptoms of TB. 40% reported they would feel ashamed of a TB diagnosis.

**Conclusions:** Most with TB familiarity knew of TB symptoms and cause, but were less informed about mode of transmission and treatment. The majority of subjects had negative attitudes towards TB. This shows a need to expand TB education in the Ngäbe-Buglé communities.

## Background

For much of the past ten years, Panama has had one of the highest tuberculosis (TB) mortality rates out of all countries in Central America (1). In 2016, the incidence rate of TB in Panama was 55 cases per 100,000 people (2). In particular, organizations providing medical services in Panama, such as Floating Doctors, have seen high rates in specific communities in the Bocas del Toro region of Panama. Lack of awareness of TB symptoms and prevention strategies is a significant risk factor for infection (3). These are compounded by a lack of knowledge of treatment options and stigma towards the disease, both of which “contribute to poor adherence and treatment compliance” (4).

To our knowledge there currently are no published studies that have investigated the TB knowledge level among residents in the Bocas del Toro area. The patients in this community have limited access to medical care, and no TB community education programs in the region exist. Furthermore, almost no epidemiological data in regards to incidence or prevalence of TB exists for these specific areas. The survey for this study was designed in conjunction with Floating Doctors, a nonprofit that has provided consistent medical care within the Bocas del Toro region for the last 5 years. Over this time period, they have observed poor treatment adherence of patients diagnosed with TB, with numerous cases noted, and little to none community knowledge in regards to the disease. Given the lack of information, the purpose of this study is to determine the baseline level of knowledge in the transmission and treatment of TB and identify cultural stigmas and other treatment obstacles residents in the Bocas del Toro region of Panama face.

Knowledge, Attitude, and Practices (KAP) surveys have been shown to be essential in establishing disease control. Rural communities of Sudan, Cameroon, and Ethiopia have comparable populations to the Bocas del Toro region and have shown strong associations between these sociodemographic factors and overall awareness of TB (3– 5). Lack of TB knowledge, negative attitudes, and improper practices have adverse effects on TB prevention and control (6–8). The analysis of these communities’ knowledge, attitudes, and practice may lead to effective community-based interventions aimed at decreasing TB prevalence and improving TB control in the Ngäbe-Buglé population of Bocas del Toro Province.

## Materials and Methods

This study was conducted in Ngäbe-Buglé communities, the indigenous population of Bocas del Toro Province, in Panama. Bocas del Toro Province is made up of mainland Panama and nine main islands. It has an area of about 430 km^2^ and a population of over 20,000 indigenous people. The structure of the healthcare system in the Ngäbe-Buglé population relies heavily on a nonprofit, mobile, medical relief organization called Floating Doctors. The Floating Doctors’ mission is to reduce the present and future burden of disease in the developing world and to promote improvements in health care delivery worldwide (9). Every week volunteers go to different remote and rural islands to provide medical care during the span of two to four consecutive days. Floating Doctors staff and volunteers then return to each community every 10 to 12 weeks to serve as the primary care providers for the community.

### Study area and population

This survey was carried out in the Ngäbe-Buglé indigenous population of Bocas del Toro Province in Panama. The Bocas del Toro Province has an area of about 430 km2 spanning the mainland of Panama in addition to nine main islands. The population of this community is approximately 20,000 people. The study was distributed in five specific communities in this region: five Ngäbe-Buglé communities: Rio Caña, Quebrada Sal, Loma Partida, Isla Tigre, and Norteño.

### Study Design and Sampling

Patients over the age of eighteen who visit the Floating Doctors free mobile clinic were selected in the waiting room before or after being seen by medical providers. Seven medical students explained the purpose of the study and received informed consent. Patients were told that participation in the study would have no effect on their medical care. Surveys were then distributed without collecting identifying information. If needed, Spanish or Ngäbe interpreters assisted in administering the consent or survey. The survey and the answer choices were read aloud, and the paper survey was filled out by the medical students by hand so that the survey was inclusive of all community members, regardless of language or literacy.

### Data Collection and Statistical Analysis

The survey was designed based on the experience and observations of long-time employees of Floating Doctors and the health disparities they noted in the perception and knowledge of TB specific to the Ngäbe-Buglé population (Figure 1). Questions were also taken from World Health Organization (WHO) guidelines to KAP for TB control and adapted for the specific population in Bocas Del Toro (10). The study instrument focused on the primary demographic data, education level, and contact with TB patients. The questionnaire developed had both open and closed-ended questions, and it was translated from English to Spanish. The survey was divided into two main sections: specific questions about TB and open-ended questions about TB related practices. If respondents indicated they did not know anything about TB or had never heard of it, they only completed the section on TB related practices. Between the two sections of the survey on Figure 1, 9 questions were in regards to TB knowledge (Figure 1,Q 6-14), 1 question on attitudes/stigma associated with TB (Figure 1, Q 15), and 5 questions on practices related to TB symptoms (Figure 1, Q 1-5).

**Figure 1:**
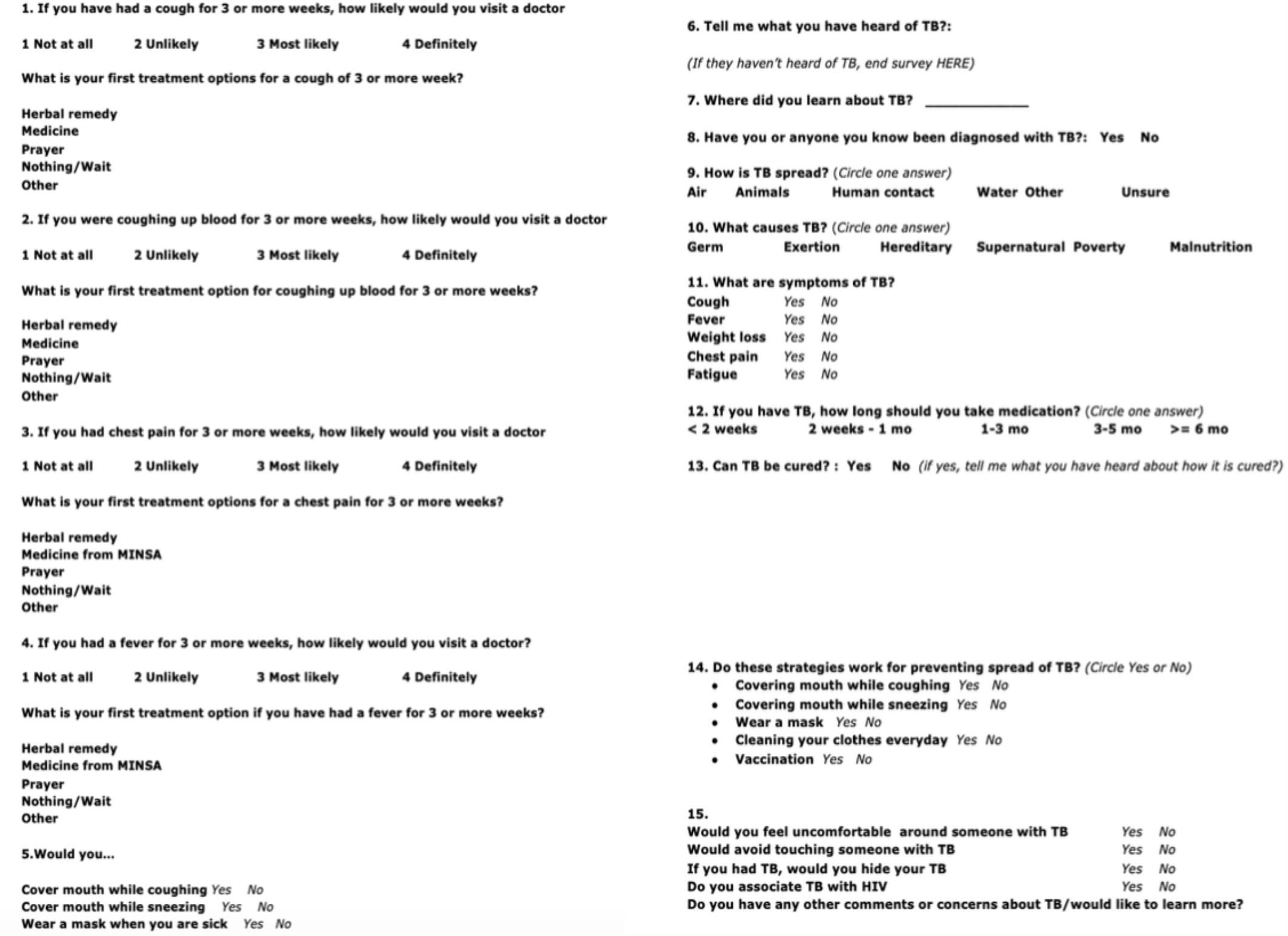
TB Survey

The questions were analyzed in Microsoft Excel individually based on the percent of individuals who answered the knowledge questions correctly as well as quantifying the percentage of answers within each category.

### Ethical considerations

The study proposal was approved by the Institutional Review Board Human Research Protections at the University of California, Irvine. Verbal informed consent was obtained from the respondents before the administration of questionnaires, and confidentiality was maintained. Age, sex, education level, and marital status information were obtained. No name, date of birth, or address was collected.

## Results

### Social Demographics of Respondents

A total of 106 participants were surveyed. Demographics of their communities and genders can be seen in Table 1.

**Table 1:** The number of participants in the survey organized into gender and community residence

|  |  |
| --- | --- |
| **Total Participants** | **106** |
| Males | 39 |
| Females | 67 |
| Average Education Level | 5<sup>th</sup> grade |
| Loma Partida Participants | 11 |
| Isla Tigre Participants | 13 |
| Rio Cana Participants | 13 |
| Norteno Participants | 39 |

### Knowledge of TB

The Knowledge questions were administered to study subjects who provided an answer to an open-ended question (Figure 2) asking about what they knew about TB. The knowledge-based portion survey included questions on the mode of TB transmission, cause, symptoms, treatment/cure, and TB prevention (Figure 1). A total of 68 (64.2%) patients expressed having some knowledge of TB. The breakdown of the percentage of respondents in each community who had knowledge of TB can also be seen in Figure 3.

**Figure 2:**
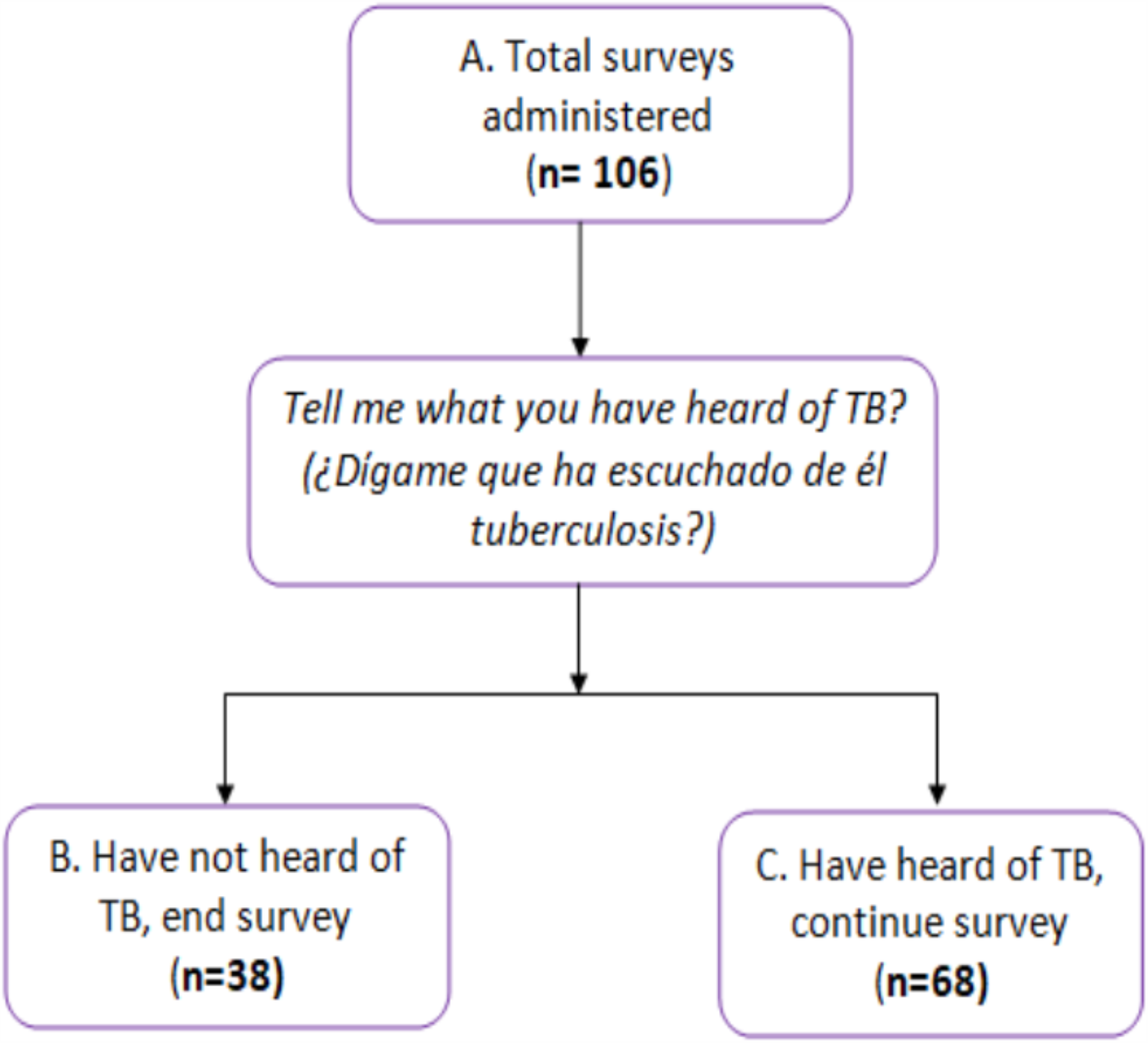
TB Knowledge

**Figure 3:**
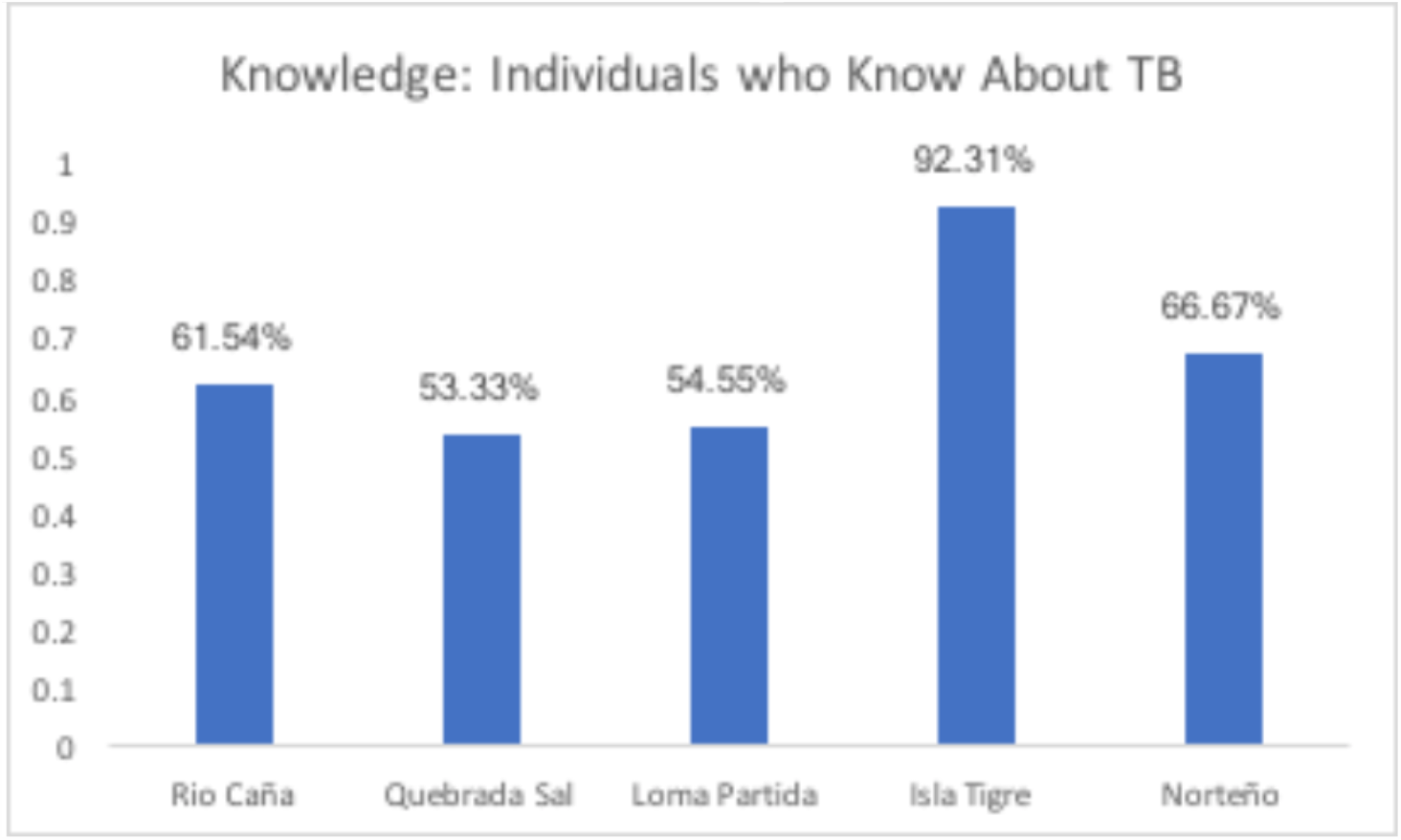
Breakdown of TB Knowledge Across Communities

Of those with knowledge of TB, 44.1% correctly answered that air is the mode of transmission for TB (Figure 4). The second more common response for the mode of transmission was human contact 23.5% of respondents. The majority of respondents, 63.2%, knew that bacteria is the cause of TB (Figure 4). Symptoms of cough (97.1%), fever (85.3%), weight loss (92.6%), chest pain (97.1%), and fatigue (80%) were correctly identified as relating to TB by the majority of patients. When asked for the appropriate duration of treatment for TB, 60% patients correctly selected more than five weeks, and 84% patients knew that TB could be cured. For preventative measures relating to TB, 86.7% patients selected covering mouth while coughing and sneezing, 81.3% correctly selected wearing a mask, 84% identified washing clothes every day, and 88% patients chose vaccines to be another form of prevention.

**Figure 4:**
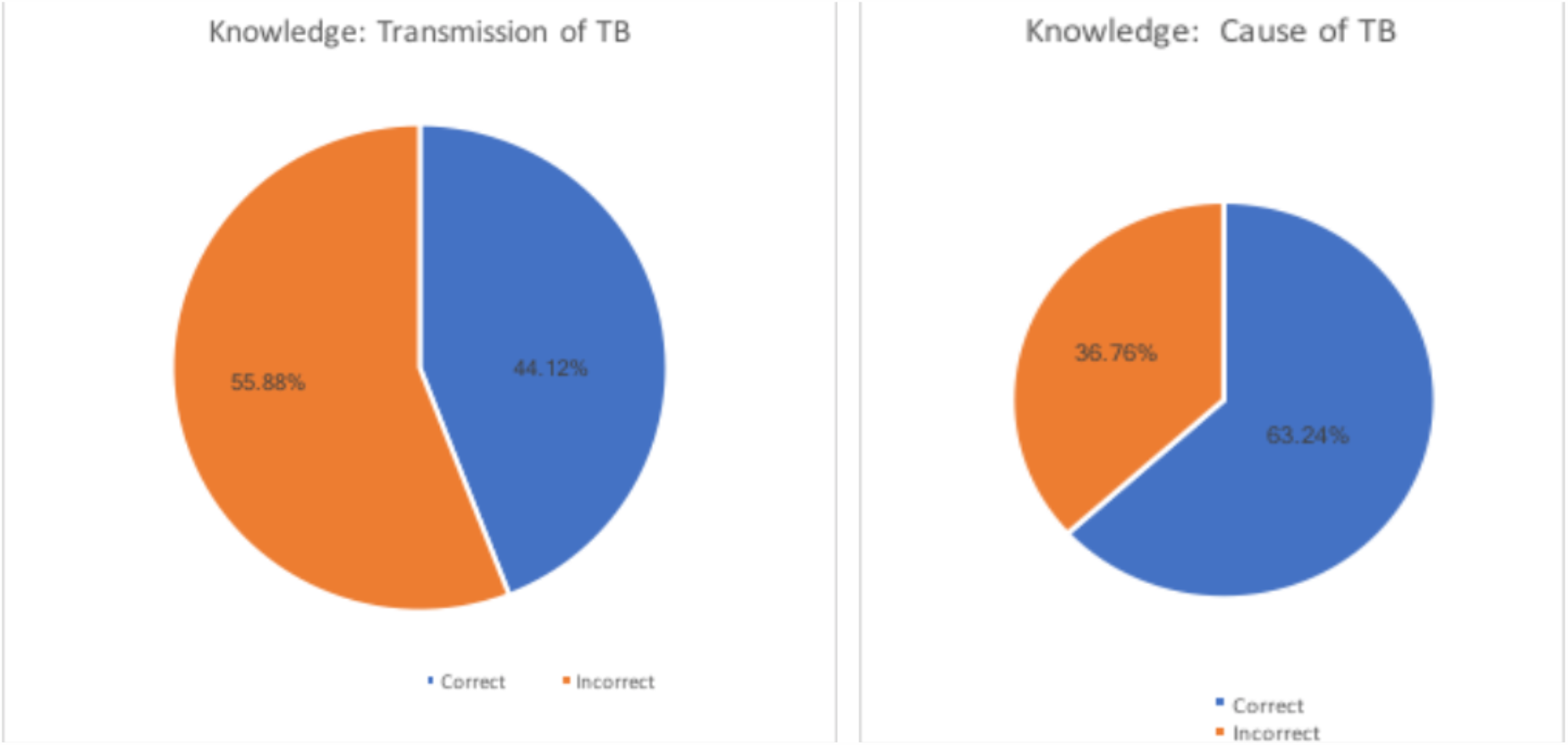
Transmissions and Causes of TB

### Attitudes of TB

Respondents’ Attitudes were evaluated based on four questions (Figure 1, Q 15). Only individuals who expressed having any knowledge of TB answered the Attitude questions. The Attitude questions asked if the individual would feel uncomfortable around TB patients, would avoid touching TB patients, would feel ashamed of a TB diagnosis, and if they associate TB with HIV. Among the 68 subjects who answered the questions, 68% said that they would be uncomfortable around TB patients, 72% would avoid touching TB patients, 40% would feel ashamed of a TB diagnosis, and 43% associate TB with HIV (Figure 5).

**Figure 5:**
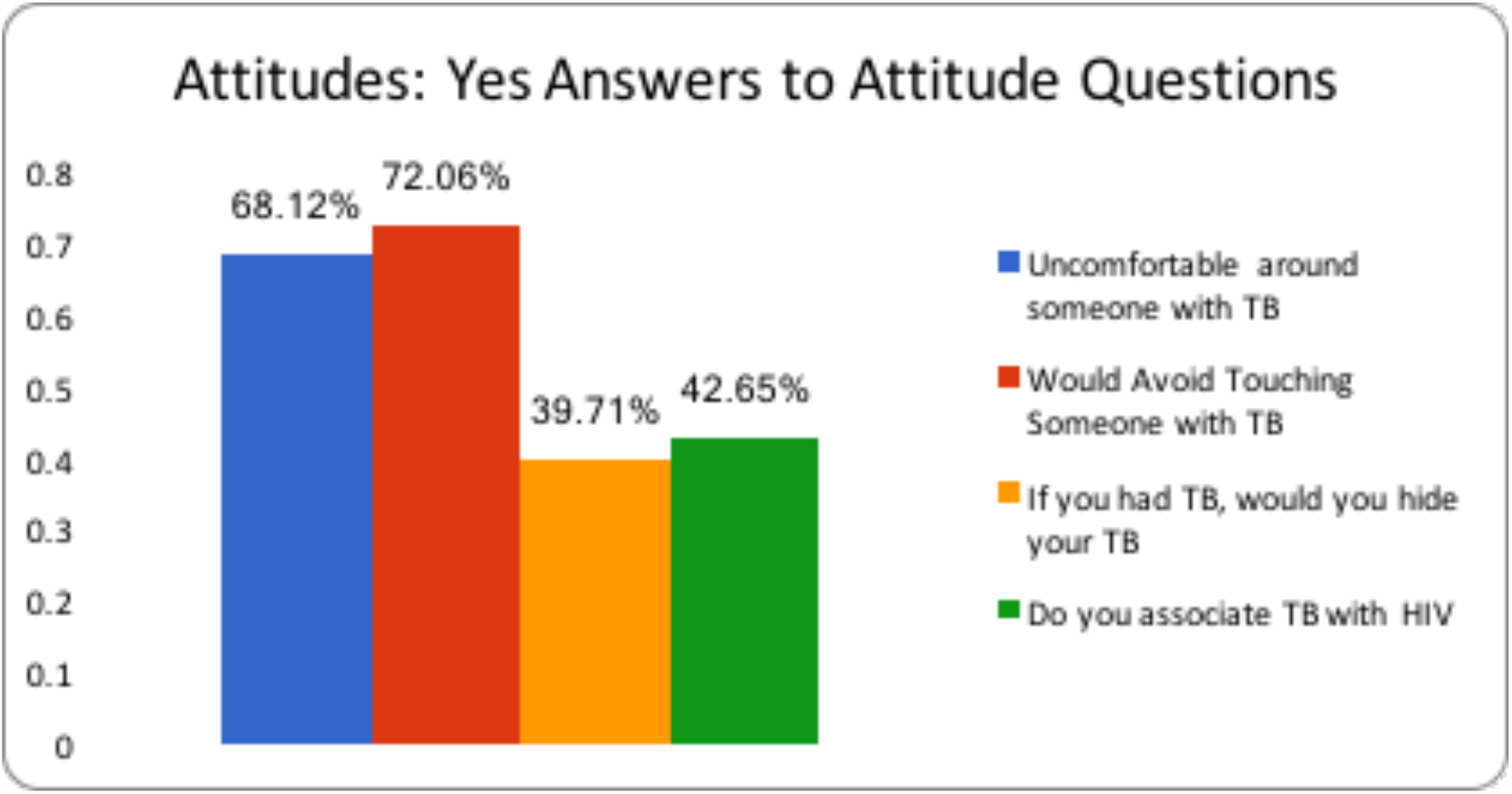
Attitudes Towards TB.

### Practices of TB

The five questions on Practices (Figure 1, Q 1-5) were asked to all 106 patients since they were included early in the survey before the knowledge questions. Medicine was the preferred method of treatment all four symptoms asked (Figure 6). Overall medicine was selected by 61.3% patients as first choice of treatment for a cough, 68.9% for cough with blood, 76.4% for chest pain and by 77.4% patients for fever of three or more weeks. The second most prevalent treatment option for all four Practice questions was herbal remedies.

**Figure 6:**
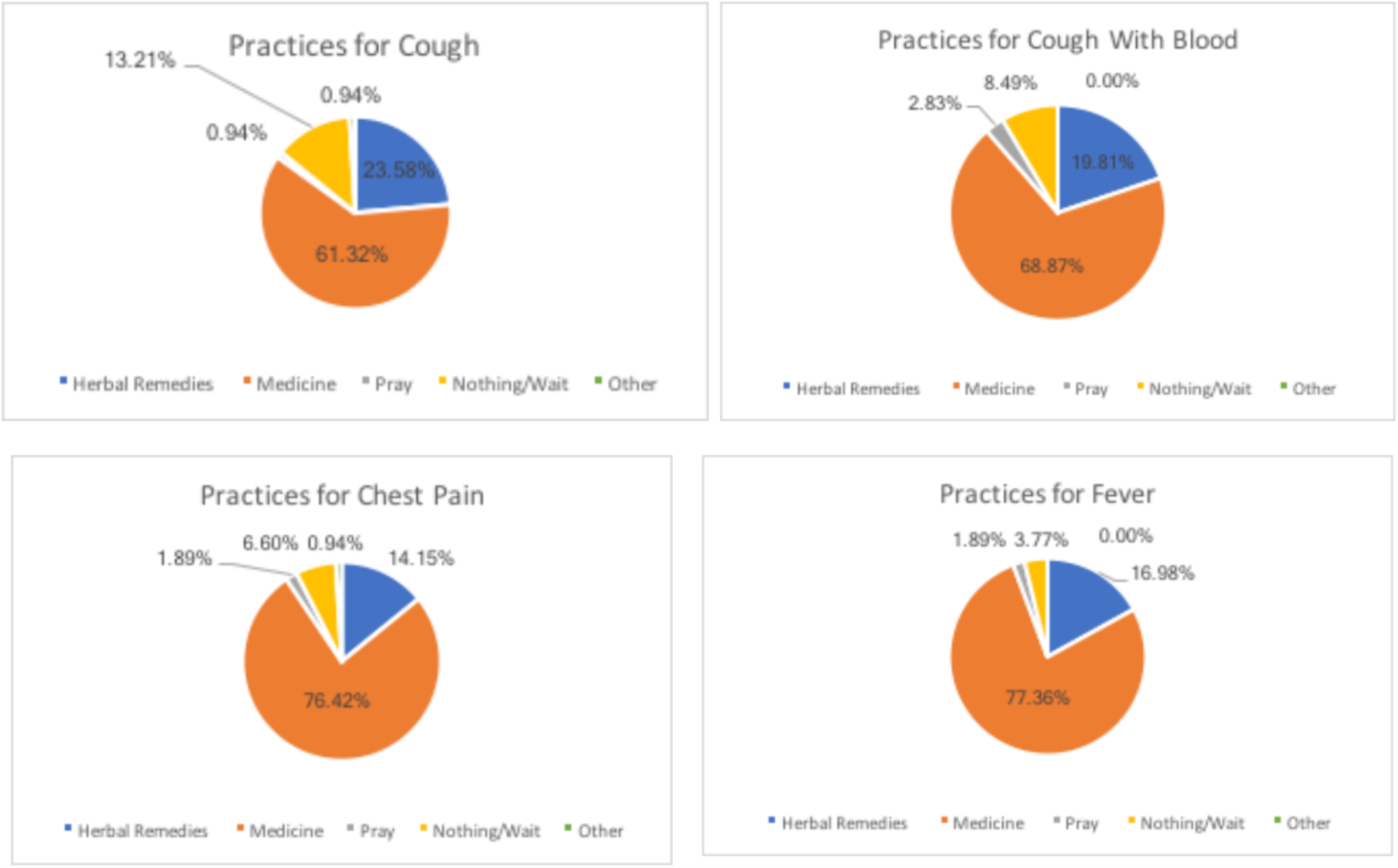
Practices Towards TB.

## Discussion

This survey was designed to establish a baseline of the knowledge, attitudes, and practices associated with TB, a severe infectious disease that is prevalent among the Ngäbe-Buglé population living in the rural communities of Bocas del Toro Province. Of the 106 patients that were surveyed, only 68 (64%) expressed hearing or knowing what TB was. However, not all TB answers given by this 64 % of patients were correct.

Focusing on the population of patients with knowledge of the disease, information in regards to the pathology of the disease and how it was treated was fairly low. Less than half of the responders knew that TB was transmitted through the air, which is crucial to preventing the spread of disease. Less than two thirds of the patients knew how long TB treatment is needed, which is also a cornerstone of preventing further disease spread. Any program designed to help educate these communities in the future should focus on targeting these two areas of knowledge gap.

The responses to the yes/no questions in regards to the symptoms seen in TB were over 80% correct in all categories (Figure 1, Q 11), indicating that patient’s do have a good idea of what symptoms are associated with TB. However, some of these symptoms are fairly general in common medical conditions, leaving the question as to how well respondents attributed them specifically to TB. Similarly, most of the patients answered with high accuracy in regards to methods to prevent the spread of TB, as well as the practices associated with perception of TB symptoms (Figure 1, Q 1-5). Overall, it seems that respondents are fairly aware of what symptoms are present in TB, but education should focus on emphasizing the length and severity of these symptoms, and helping them differentiate TB from other common medical conditions such as the common cold, flu, pneumonia, etc.

Interestingly, there is a differing proportion of TB knowledge within these five communities. Of the patients surveyed, 92.3% in Isla Tigre reported that they knew about TB, significantly higher than any other community. Patients expressed multiple sources for where they first heard about TB varying from a family member, neighbor, or from witnessing it directly. It could very well be possible that the higher percentage of knowledge could related to a higher prevalence of TB in this specific community, though further research would be needed to investigate this hypothesis.

Another area of concern that was brought up by the survey was the attitudes in regards to the disease. Given the high percentage of respondents who noted they would feel uncomfortable around individuals with TB, and ashamed with the diagnosis, a serious issue exists in the sociocultural norms associated with the disease in the region. Further research and emphasis needs to be placed to investigate what exact aspects of the disease stigmatize people.

Considering that all these questions were asked in the survey, a study limitation exists that some of the correct responses we received for such knowledge categories were answered correctly by chance. Therefore, there may be a more significant deficit in TB knowledge. A more detailed survey with more open ended questions would help address this issue.

## Conclusions

Our study is the first of its kind to demonstrate that the Ngäbe-Buglé communities of Bocas del Toro Province do not fully understand TB and that there is a need for TB education in this population. Education specifically on TB spread, treatments, and differentiating it from other medical conditions is needed in all communities. Additionally, an educational program will help surmount the negative attitudes and connotations surrounding TB, so that community members feel comfortable seeking medical attention if TB symptoms arise. Bocas del Toro Province would benefit from future studies that investigate these differences to identify specific steps to take for improving TB knowledge, attitudes, and practices. We hope this study serves as groundwork to help expand health education for the Ngäbe-Buglé communities in Bocas del Toro, Panama.

## Data Availability

All data in the manuscript can be requested from the authors.

## Authors’ Statements

### Author Contributions

LRC, KE, SR, EF, LC, PB, AC, EC conceived the study; LC, KE, SR, EF, LC, PB, AC, EC designed the study protocol; LRC, KE, SR, EF, LC, PB, AC, EC carried out the survey assessment; LRC, KE, SR, EF, LC, PB, AC, EC carried out the analysis and interpretation of these data LRC, KE, SR, EF, LC, PB, AC, EC drafted the manuscript; EF, KE, EC, PB critically revised the manuscript for intellectual content. All authors read and approved the final manuscript. PB is the guarantor of the paper.

## Acknowledgements

We would like to acknowledge Floating Doctors for providing the clinic organization and transportation in Bocas del Toro. We are grateful to have partnered with this non-profit organization.

## Ethical Approval

This is an original manuscript, unpublished, and not under consideration elsewhere.

## References

1. Tarajia M, Goodridge A. Tuberculosis remains a challenge despite economic growth in Panama. Int J Tuberc Lung Dis Off J Int Union Tuberc Lung Dis. 2014 Mar;18(3):286–8.

2. Panama Tuberculosis Profile [Internet]. Geneva, Switzerland: World Health Organization; 2019 Oct. Available from: https://extranet.who.int/sree/Reports?op=Replet&name=%2FWHO_HQ_Reports%2FG2%2FPROD%2FEXT%2FTBCountryProfile&ISO2=PA&LAN=EN&outtype=pdf

3. Suleiman MMA, Sahal N, Sodemann M, Elsony A, Aro AR. Tuberculosis awareness in Gezira, Sudan: knowledge, attitude and practice case-control survey. East Mediterr Health J Rev Sante Mediterr Orient Al-Majallah Al-Sihhiyah Li-Sharq Al-Mutawassit. 2014 Mar 13;20(2):120–9.

4. Esmael A, Ali I, Agonafir M, Desale A, Yaregal Z, Desta K. Assessment of patients’ knowledge, attitude, and practice regarding pulmonary tuberculosis in eastern Amhara regional state, Ethiopia: cross-sectional study. Am J Trop Med Hyg. 2013 Apr;88(4):785–8.

5. Kwedi Nolna S, Kammogne ID, Ndzinga R, Afanda B, Ntonè R, Boum Y, et al. Community knowledge, attitudes and practices in relation to tuberculosis in Cameroon. Int J Tuberc Lung Dis Off J Int Union Tuberc Lung Dis. 2016;20(9):1199–204.

6. Balogun M, Sekoni A, Meloni ST, Odukoya O, Onajole A, Longe-Peters O, et al. Trained community volunteers improve tuberculosis knowledge and attitudes among adults in a periurban community in southwest Nigeria. Am J Trop Med Hyg. 2015 Mar;92(3):625–32.

7. Anochie PI, Onyeneke EC, Onyeozirila AC, Igbolekwu LC, Onyeneke BC, Ogu AC. Evaluation of public awareness and attitude to pulmonary tuberculosis in a Nigerian rural community. Germs. 2013 Jun 1;3(2):52–62.

8. Tobin EA, Okojie P-W, Isah EC. Community knowledge and attitude to pulmonary tuberculosis in rural Edo state, Nigeria. Ann Afr Med. 2013 Sep;12(3):148– 54.

9. Our Mission [Internet]. Floating Doctors; Available from: https://floatingdoctors.com/about-us-who-we-are/our-mission/

10. Advocacy, communication and social mobilization for TB control: A Guide to Developing Knowledge, Attitude, and Practice Surveys [Internet]. Geneva, Switzerland: World Health Organization; 2018. Available from: https://apps.who.int/iris/bitstream/handle/10665/43790/9789241596176_eng.pdf?sequence=1

